# Small but systematic bias introduced by EEG electrodes in PET imaging

**DOI:** 10.64898/2026.08.12.26360268

**Authors:** P Stöhrmann, M Ponce de León, G Dörl, C Milz, S Graf, B Eggerstorfer, MB Reed, M Murgaš, P Falb, K Al Barede, L Nics, S Rasul, M Hacker, R Lanzenberger, A Hahn

## Abstract

**Purpose:** Attenuation correction (AC) of PET images is essential for accurate quantification. Brain PET studies comprising simultaneous EEG (PET_EEG_) may suffer from metal artifacts in CT images (CT_EEG_), or improper correction when electrodes are not present in the CT (CT_0_). As these influences are not well-characterized, we aim to compare metal artifact reduction (MAR) techniques for CT_EEG_ images, and evaluate differences between attenuated-corrected PET_EEG_ using CT_0_ and CT_EEG_ with MAR, synthetically placed electrodes (CT_EEG-synth_) and extended Hounsfield unit (HU) range.

**Methods:** 19 healthy participants underwent two total-body PET/CT scans with [^18^F]FDG, with and without 32 EEG scalp electrodes, respectively. We evaluated five MARs to reduce streaks caused by the EEG electrodes in the CT_EEG_. Finally, CT_0_, CT_EEG_ with (CT_EEG-iMAR-Ext_) and without extended HU range (CT_EEG-iMAR_) and CT_EEG-synth_ were used to perform attenuation correction of PET_EEG_. We compared our results to PET_0_/CT_0_ scan using relative differences.

**Results:** CT_EEG_ and CT_EEG-iMAR_ showed the smallest differences to CT_0_. PET_EEG_/CT_EEG-iMAR-Ext_ exhibited the lowest differences to PET_0_/CT_0_ (average bias across all regions of −0.46%), followed by similar performance of PET_EEG_/CT_EEG-iMAR_ (−0.73%) and PET_EEG_/CT_EEG_ (−0.76%). Conversely, PET_EEG_/CT_0_ demonstrated the largest average differences (−1.81%), with values reaching −2.71% in the parietal lobe. These differences were consistent across subjects, yielding significant effects in most of the brain (p_FWE_ < 0.05). CT_EEG-synth_ performed not as good as CT_EEG_ (−1.21%).

**Conclusions:** CT_EEG_ with extended HU range is most suitable for attenuation correction of PET_EEG_ images, with MAR correction offering little additional improvement.

## Introduction

The development of multimodal neuroimaging techniques offers several advantages for studying the nervous system under normal and pathological conditions. PET/CT scanners allow for the nearly simultaneous acquisition of anatomical and functional data without moving the patient between scanners, thus avoiding image misregistration. The additional integration of EEG recordings provides information with high temporal resolution in the order of milliseconds, thus complementing PET frames, which offer better spatial localization but are rarely shorter than seconds, despite recent advances in image reconstruction and total-body scanners.

PET/CT measurements may be affected by patients’ dental implants/fillings, prostheses, and cardiac pacemakers [1]. These often contain metals, which have high atomic numbers and are radiologically much denser than human tissue. As a result, bright and dark streak artifacts appear on CT images, not only at the location of the metallic object but also in the surrounding area, with the effect being more pronounced in the case of low-dose CT [2, 3]. This affects not only the CT image but also the attenuation-corrected PET image [4], though the effects are not as easily discernible [5]. Nonetheless, they may lead to an over- or underestimation of tracer uptake, which could adversely affect image interpretation and quantification and thus the patient’s diagnosis [6]. However, the impact of these artifacts on the quantification of attenuation-corrected brain PET images has not been studied in depth. The existing literature on PET analysis with artifacts in CTs is extensive, but focuses on prostheses, whose effect depend on their position and size, factors which differ from those of EEG electrodes [7, 8].

PET images must be corrected for attenuation and scatter using transmission scans or attenuation maps derived from CT scans, or via attenuation maps extrapolated from MRIs in the case of PET/MR scanners. As standalone PET scanners with transmission sources become less common, transmission scans are increasingly being replaced by CT scans. Thus, previous research has focused on transforming Hounsfield units (HU) measured by CT scanners at different X-ray tube voltages into the 511 keV linear attenuation maps for attenuation correction of PET data [9]. However, information on linear attenuation coefficients (LAC) at values above 2000 HU is limited [10, 11], whereas metals markedly exceed this limit [12–14]. It is important to note, that most of today’s PET/CT scanners store CT-transmission scans as 12-bit data, as opposed to more advanced scanners that can store 16-bit data. The former allows values ranging from −1,024 to 3,071 HU ([-1024, −1024 + 2^12^) at 1 HU increments (ΔHU_min_ = 1)), limiting their ability to encode the true HU values of and differentiate between high-density materials. On the other hand, 16-bit scanners allow values ranging from −32,768 to 32,767 HU. A third option makes use of the image header attribute “multiplicative scaling”, to store larger values in extended 12-bit format ([-10240, −10240 + 10·2^12^)) at the cost of coarser HU resolution (ΔHU_min_ = 10) [15].

While appropriate data formats solve the issue of artificially capped HU, image artefacts resulting from metals in the FOV are unaffected by this. Several commercial and open-source metal artifact reduction (MAR) techniques have been proposed in order to reduce artifacts in CT images, but there is still no standard reference. This remains one of the major challenges in PET/CT imaging with metal artifacts. The iterative MAR (iMAR) algorithm is a commercial approach offered by Siemens Healthineers for their PET/CT scanners. It has been shown to significantly reduce metal artifacts caused by prostheses and dental implants [16]. The advantage of this approach is that attenuation-corrected PET images can be obtained directly from the scanner, without the need for in-house image-processing expertise. Free software based on the metal deletion technique (MDT) is also available (www.revisionrads.com/)[17]. Previous research has shown that the MDT is more effective than the commercial iMAR for dental implants, but information on EEG electrodes is not yet available to our knowledge [18]. Other possibilities are the refined MAR (RMAR) and the normalized MAR (NMAR) provided by the wxDicom 2.61 software [19]. All of the aforementioned techniques have been tested mainly on thoracic and pelvic images, but information on skull and brain imaging with EEG is limited.

This study was divided into two main objectives to obtain proper brain PET_EEG_ images (i.e., PET scans with EEG-electrodes present) compared to the reference (PET_0_ corrected for attenuation using CT_0_, i.e., both without EEG): The first goal was to evaluate the performance of different MAR techniques on CT_EEG_ images (CT_EEG-MAR_) to find the one with the smallest differences to the CT_0_ image. Secondly, we compared the PET_EEG_ image corrected for attenuation using either a) CT_EEG_, b) CT_EEG-MAR_ identified in objective 1 (with default HU ranges), c) CT_EEG-MAR-Ext_ (with extended HU ranges), d) CT_EEG-synth_ (synthetic incorporation of EEG electrodes in CT_0_ images to simulate a possible compensation for the lack of this information, e.g., for pseudo-CTs from PET/MR scans), and e) CT_0_ to the reference.

## Materials and methods

### Participants and study design

21 healthy participants (mean age = 24.52 ± 4.96 years, 13 females) underwent a 105-min whole-body PET/CT scan with bolus plus continuous infusion of the radiotracer [^18^F]FDG. First, a CT scan was performed with the EEG cap (CT_EEG_). Simultaneously, PET and EEG (PET_EEG_) were acquired with the subjects’ eyes open (20 min) and then with their eyes closed (70 min). The EEG cap was removed thereafter and a CT_0_ and a 10-min PET_0_ scan were performed (see also figure 1). During recording, light was dimmed and noise kept to a minimum.

**Fig. 1:**
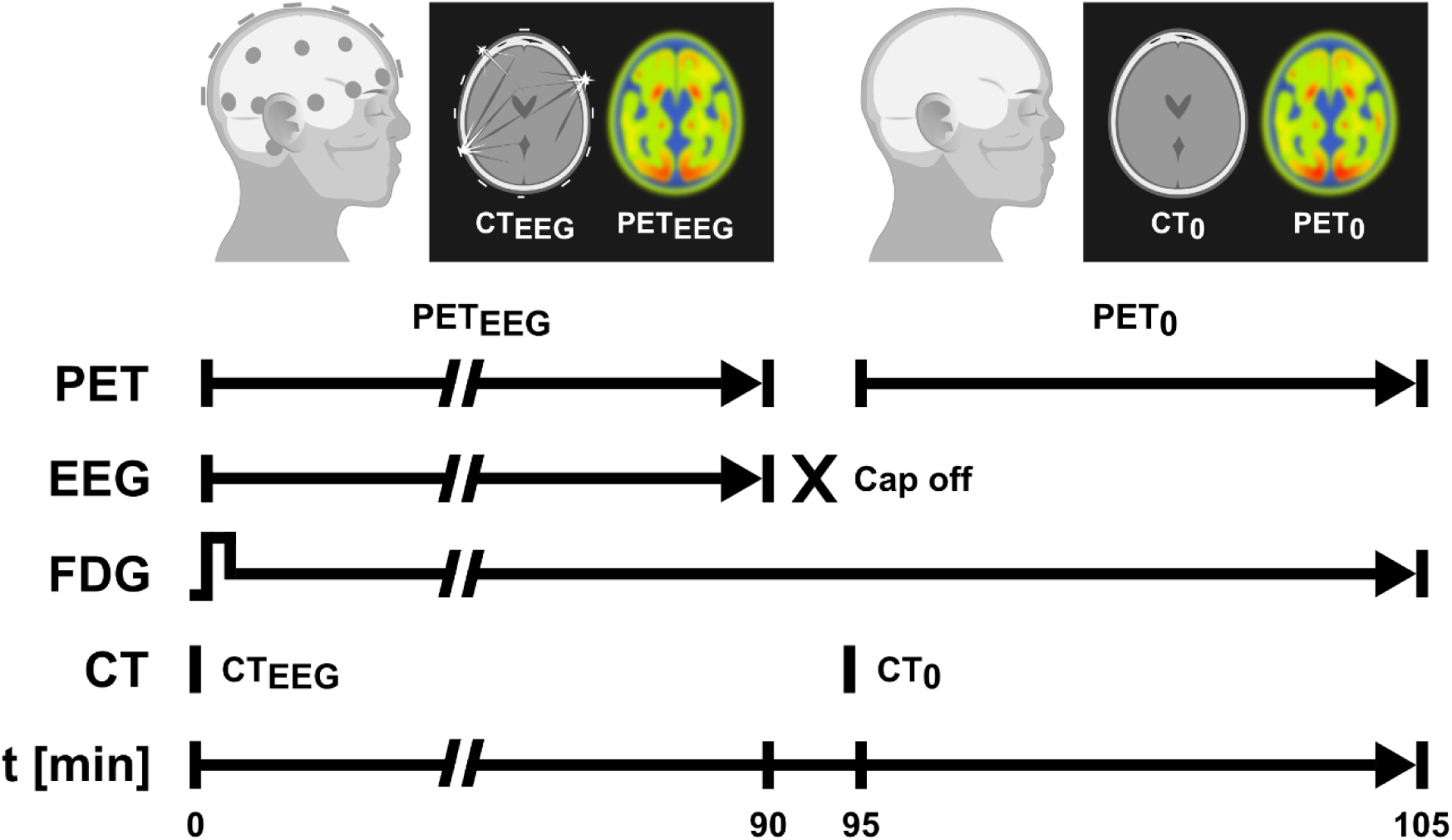
Schematic description of the study design. CT and PET were recorded consecutively, once while the study participants were wearing a 32-lead EEG cap (CT_EEG_ and PET_EEG_, for 90 min), and once without the EEG cap (CT_0_ and PET_0_, 10 min). For the analysis, only adjacent mean PET images of 10 minutes each were used, i.e., 80-90 and ≈ 95-105 min after start of tracer application. Electrodes were well-visible in the CT_EEG,_ along with corresponding streaking artifacts. Differences between PET images were not visually discernible. The timeline is not to scale, interrupted lines represent continuation of the ongoing procedure. This figure was created in Inkscape.

All participants underwent a standard medical examination performed by an experienced psychiatrist at the initial screening visit, including blood tests, electrocardiography, neurological testing and the Structured Clinical Interview for DSM-IV. Female participants also underwent a urine pregnancy test during the screening visit and upon arrival on the day of the PET/CT scan. Exclusion criteria included current and past (12 months) somatic, neurological, or psychiatric disorders; current and past substance abuse or psychotropic medication; current pregnancy or lactation; and prior research-related radiation exposure in the past 10 years. All participants gave written informed consent and were financially compensated for their participation. The study was approved by the Ethics Committee of the Medical University of Vienna (Ethics Number: 1642/2022) and procedures were performed in accordance with the Declaration of Helsinki.

### Radiotracer application

The tracer administration was performed as bolus plus constant infusion with 5.1 MBq/kg in 50 ml physiological sodium chloride solution at a rate of 816 ml/h for 1 min and 21 ml/h for 104 min. The radiotracer was injected via the cubital vein using a perfusion pump (Syramed μSP6000, Arcomed, Regensdorf, Switzerland) and was kept in a custom wolfram shield to minimize radiation exposure.

### Data acquisition and reconstruction

PET/CT data were acquired on a Siemens Biograph Vision Quadra (Siemens Healthineers, Erlangen, Germany) (axial field-of-view: 106 cm, effective sensitivity: 2182 cps/kBq, time-of-flight resolution: 228 ps). Ultra-low-dose CT scans (140 kVp tube voltage, tube current 20 mA) were acquired and reconstructed using the regular and extended 12-bit HU range. CT images were reconstructed to a 512 × 512 × 708 image matrix with a voxel size of 1.52 × 1.52 × 1.50 mm. All PET data were reconstructed offsite using a 3D-TOF OP-OSEM algorithm with 2 iterations, 5 subsets and 4mm FWHM post-reconstruction filtering (Hann) to a matrix of 220 × 220 × 645 with a voxel size of 3.30 × 3.30 × 1.65 mm (frames: 2 × 10 min, 80-90 and ≈95-105 min after initial tracer application).

Structural MRI of the brain was acquired additionally on a Siemens Prisma 3T MR system using a T1-weighted MPRAGE sequence (echo-time/repetition-time = 2.37/1800 ms, inversion-time = 900 ms, acquisition-time = 219 s, flip angle = 8°, matrix size = 288 × 288, 208 slices, voxel size = 0.87 × 0.87 × 0.85 mm), which was used for spatial normalization. For two participants, the T1 data was unavailable; therefore, normalization was conducted utilizing PET data only.

The EEG was recorded using a 32-channel BrainCap MR (Brain Products GmbH, Gilching, Germany) with electrodes placed according to the extended international 10-20 system. Impedances were kept below 10 kΩ and the sampling rate was 5 kHz. Battery and amplifier were placed outside of the PET/CT bore and only connected to the cap during PET_EEG_. The apical cable bundle and connector box being part of the cap were fixed to the scanner bed prior to CT_EEG_.

Venous blood samples were collected at 4, 90, and 105 min. The activity of plasma and whole blood was measured using a γ-counter (Wizward2, 3”, Perkin Elmer) which had been cross-calibrated to the PET/CT scanner.

### Image processing

All image pre-processing was carried out in MATLAB R2018b (The MathWorks Inc., Massachusetts) and with SPM12 software. For this study, we analysed only the last 10 minutes of PET_EEG_ (minutes 80-90 after the start of the radiotracer application) and the 10-minute PET_0_ scan (roughly minutes 95-105 since the radiotracer application). Both CT_EEG_ and CT_0_ were used for PET reconstruction, co-registered to the respective PET sum image. The analysis at the volume of interest (VOI) level was conducted using the Harvard Oxford Atlas [20].

#### Application of MAR on CTEEG

In the present study, 5 metal artifact reduction (MAR) techniques were evaluated to reduce streaks caused by EEG electrodes in the CT images. First, we used the iMAR provided by Siemens for some of their PET/CT and also SPECT/CT scanners (Biograph Vision/mCT/Horizon and Symbia Intevo Bold). In addition, we evaluated the freely available software based on the MDT (www.revisionrads.com/)[17]. Subsequently, the refined MAR (RMAR) and the normalized MAR (NMAR) provided by the wxDicom 2.61 software [19] were performed. In addition, a 2-D adaptive noise-removal filtering (ANRF) implemented in MATLAB was investigated. The 5 MAR techniques were applied to the CT_EEG_ images.

CT processing included head extraction from whole-body scans, co-registration of the T1w MRI and the CT_0_ to the CT_EEG_, normalization of the co-registered T1w image to MNI space and inverse transformation of the atlas to the participant space. HU were averaged across VOIs.

#### Attenuation correction of PET images and processing

Initial processing included head extraction from all the whole-body CT and PET images (CT_0_, PET_0_, CT_EEG_ and PET_EEG_), co-registration of CT_0_ to PET_0_, and co-registration of CT_0_, CT_EEG_ and CT_EEG-MAR_ (MAR selected in objective 1, with and without extended HU range) to PET_EEG_. The transformation matrix was applied to the whole-body CT images. Voxels containing electrodes and cables (arbitrary threshold of ≥ 800 HU) in CT_EEG_ co-registered to CT_0_, were copied to CT_0_ to derive an additional, synthetic CT scan (CT_EEG-synth_), lacking artefacts but containing less extensive attenuation information on the EEG cap. We then performed attenuation correction of PET_0_ (with CT_0_) and PET_EEG_ (with CT_0_, CT_EEG_, CT_EEG-MAR_, CT_EEG-MAR-Ext_, and CT_EEG-synth_) images.

After attenuation correction, six whole-body PET images per subject were obtained (five for PET_EEG_ and one for PET_0_, see Table 2) and their processing included: head extraction, co-registration of all PET images to T1 MRI, and normalization to MNI space. Static PET images were normalized to the cerebellum (excluding the vermis), yielding standard uptake value ratio (SUVR), and then smoothed with an 8 mm^3^ Gaussian kernel.

### Statistical analysis

To identify which MAR technique yielded CT_EEG-MAR_ images with the smallest intracranial HU-differences compared to the reference (CT_0_), we estimated the voxel-wise relative differences of the HU between the reference and each of the five MAR techniques (CT_EEG-MAR_), alongside the comparison with CT_EEG_. The differences were averaged across VOIs and subjects.

Subsequently, we conducted a comparative analysis of PET_EEG_, corrected for attenuation using CT_EEG_, CT_EEG-MAR_, CT_EEG-MAR-Ext_, CT_EEG-synth_ and CT_0_, against the reference (PET_0_ corrected for attenuation using CT_0_). Relative voxel-wise differences were calculated between PET_EEG_ reconstructions and the reference, subsequently averaging these differences within VOIs and across subjects.

Finally, we compared PET_EEG_, corrected for attenuation using the best performing MAR approach as determined in objective 1, with (CT_EEG-iMAR-Ext_) and without extended HU range (CT_EEG-iMAR_), with the reference (PET_0_ corrected for attenuation using CT_0_) (objective 2). As with our initial objective, voxel-wise relative differences were calculated and averaged. Furthermore, two-sided paired t-tests were performed in SPM to identify brain regions in PET_EEG_ scans whose SUVR is significantly affected by changing the AC scan from CT_EEG_ to CT_0_ (as in the case when either only CT_0_ is available or a pseudo-CT was derived from an MRI) or CT_EEG_ to CT_EEG-iMAR-ext_.

## Results

Data from 19 participants (25.11 ± 5.15 years; 12 females) were included in the analysis. One subject was excluded from the final analysis due to an aborted image acquisition, and a second one due to an incidental finding of an anatomical anomaly without previous clinical symptoms.

### Electrode attenuation in CT

In the unprocessed CT scans without extended HU range, image intensity was capped at 3071 HU. Extending the HU range lead to intra-scan maximum values of roughly 14,000 to 17,000 HU for the used Ag/AgCl electrodes and above 25,000 HU in two subjects with dental implants. Applying the conversion method proposed by Oehmigen et al. [10] one ends up with a LAC in the range of approximately 0.33-0.38 cm^-1^. The x-ray mass attenuation coefficient of elemental silver and total attenuation with coherent scattering of AgCl in the NIST Standard Reference Databases 8 and 126 for 0.5 MeV are given as μ_en_/ρ = 3.822·10^-2^ cm^2^/g (≈ 0.4 cm^-1^) and 9.106·10^-2^ cm^2^/g (≈ 0.5 cm^-1^), respectively [21, 22]. In their investigation on material identification of foreign bodies with CT, Bolliger and colleagues reported silver having a mean HU value of 16,949 [14].

### Performance of MAR approaches on CT

Comparisons between CT_0_ and CT_EEG_, as well as the five MAR approaches (CT_iMAR_, CT_MDT_, CT_RMAR_, CT_NMAR_, and CT_ANRF_) are shown in Table 1 and Figure 2. iMAR was the metal artefact removal technique that demonstrated the most comparable HU on average (4.02 ± 4.96 %), and thus small relative differences across all VOIs in comparison to CT_0_. Although CT_EEG-iMAR_ performed best on both average and in superficial brain regions, subcortical regions showed more bias than native CT_EEG_. Conversely, RMAR exhibited higher differences in all VOIs. MDT, NMAR and ANRF techniques showed intermediate performance across VOIs (5.22 ± 6.18, −10.33 ± 4.52, 16.12 ± 5.42, respectively). MDT and ANRF exhibited highly positive and in absolute values larger numbers in cortical areas but smaller, negative values in subcortical regions, whereas NMAR showed only negative values, which were largest in subcortical regions. Only the iMAR approach was further evaluated for its effects on PET attenuation correction.

**Fig. 2:**
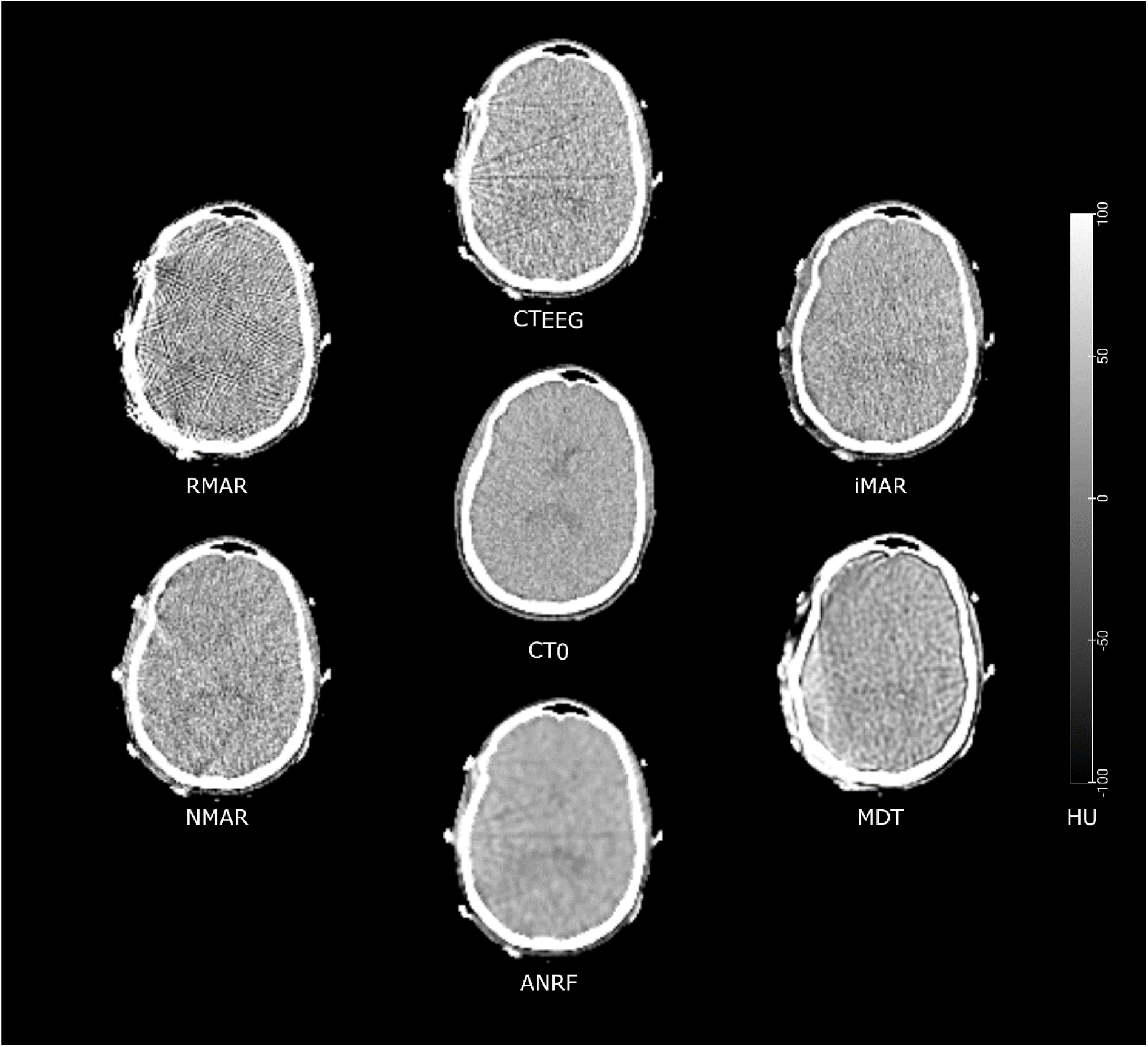
CT images of a representative participant showing the comparison of the different metal artifact reduction (MAR) techniques. CT_0_ (centre) refers to the reference standard (i.e., CT acquisition without EEG) and CT_EEG_ (top) to acquisition with EEG. The five CTs corrected with MAR are shown in the following order (clock-wise): vendor-provided MAR (iMAR), metal deletion software (MDT), adaptive noise removal filtering (ANRF), as well as refined (RMAR) and normalized MAR (NMAR). Image windowing is identical across methods and narrowly windowed ([-100,100] HU) to enhance visibility of artifacts. In combination with low-dose CT, this explains the lack of anatomical, soft-tissue contrast. Note, how MAR techniques differently affect smoothness, brightness and structure of signal and noise. Data visualization with MRIcroGL, arranged in GIMP.

**Table 1:** Percentage difference of Hounsfield units for different CT acquisitions and metal artifact reduction (MAR) techniques (CT_EEG-iMAR_, CT_EEG-MDT_, CT_EEG-RMAR_, CT_EEG-NMAR_ and CT_EEG-ANRF_). All comparisons were performed with respect to the reference (CT_0_, acquisition without EEG). Values correspond to mean ± standard deviation (averaged across subjects and volumes of interest). Abbreviations: iMAR: iterative MAR as provided by the vendor, MDT: metal deletion technique, RMAR: refined MAR, NMAR: normalized MAR (NMAR), ANRF: 2-D adaptive noise-removal filtering. **Bold** and *italic* show the least and second to least absolute difference within each VOI, respectively.

EEG influence on PET
| Region | CT<sub>0</sub> |  |  |  |  |  |
| --- | --- | --- | --- | --- | --- | --- |
|  | vs |  |  |  |  |  |
|  | CT<sub>EEG</sub><br>[%] | CT<sub>EEG-iMAR</sub><br>[%] | CT<sub>EEG-MDT</sub><br>[%] | CT<sub>EEG-RMAR</sub><br>[%] | CT<sub>EEG-NMAR</sub><br>[%] | CT<sub>EEG-ANRF</sub><br>[%] |
| Frontal | 8.17 ± 2.08 | **-3.74 ± 4.29** | 12.16 ± 3.41 | -16.80 ± 5.60 | -9.36 ± 2.18 | 35.96 ± 4.98 |
| Occipital | 7.23 ± 3.30 | **-0.85 ± 4.21** | 19.31 ± 13.42 | -11.75 ± 9.73 | -6.70 ± 4.62 | 34.57 ± 7.77 |
| Parietal | 8.36 ± 3.71 | **-5.72 ± 5.98** | 21.69 ± 7.90 | -19.45 ± 6.95 | -10.98 ± 3.44 | 34.80 ± 5.93 |
| Temporal | 7.05 ± 4.33 | 3.27 ± 4.70 | 6.88 ± 5.25 | -16.30 ± 9.23 | **-3.10 ± 5.70** | 31.94 ± 6.81 |
| Insula | **-0.36 ± 4.88** | -6.65 ± 6.17 | -4.62 ± 5.18 | -32.51 ± 7.57 | -11.20 ± 5.76 | *-1.01 ± 4.46* |
| Caudate | **-1.24 ± 4.50** | -6.41 ± 5.04 | -4.88 ± 5.40 | -34.92 ± 6.20 | -16.51 ± 3.91 | *-2.59 ± 4.97* |
| Putamen | **-1.03 ± 3.75** | -5.10 ± 4.52 | -3.38 ± 4.38 | -30.13 ± 9.84 | -11.80 ± 5.55 | *-1.39 ± 3.83* |
| Thalamus | **-3.00 ± 4.52** | -6.92 ± 4.73 | -5.42 ± 4.49 | -33.15 ± 6.30 | -12.97 ± 4.96 | *-3.36 ± 4.60* |
| Mean | **3.15 ± 3.88** | *-4.02 ± 4.96* | 5.22 ± 6.18 | -24.38 ± 7.68 | -10.33 ± 4.52 | 16.12 ± 5.42 |

In terms of time consumption, the CT_EEG-iMAR_ was obtained immediately from the scanner. The other four MAR techniques required the following times for CT enhancement: ∼50 min (CT_EEG-MDT_), ∼5 min (CT_EEG-RMAR_), ∼5 min (CT_EEG-NMAR_), and << 1 min (CT_EEG-ANRF_).

### Attenuation correction of PETEEG with different CT approaches

Table 2 and Figure 3a-c show the percentage differences between the PET_EEG_ when corrected for attenuation using the different CT maps (CT_0_, CT_EEG_, CT_EEG-iMAR_, CT_EEG-iMAR-Ext_ and CT_EEG-synth_) and the reference (PET_0_/CT_0_). The PET_EEG_/CT_EEG_ (mean ± SD = −0.76 ± 2.14 %) and CT_EEG-iMAR_ (−0.73 ± 2.14 %) exhibited little bias compared to the PET_0_/CT_0_, but the lowest differences were observed for the reconstruction with extended HU range, i.e., PET_EEG_/CT_EEG-iMAR-Ext_ (−0.46 ± 2.16 %). Attenuation correction without accounting for EEG electrodes (PET_EEG_/CT_0_) showed the highest deviation from the reference (−1.81 ± 2.03 %). Synthetic incorporation of electrodes into the reference attenuation correction map (PET_EEG_/CT_EEG-synth_) showed performance between PET_EEG_/CT_EEG_ and PET_EEG_/CT_0_. Finally, we evaluated whether the temporal difference in acquisition between the reference (PET_0_/CT_0_) and the various AC approaches (PET_EEG_/CT_X_) influences our comparisons. Here, the assessment of PET_EEG_/CT_0_ vs. PET_EEG_/CT_EEG_ yielded virtually identical results as PET_EEG_/CT_EEG_ - PET_EEG_/CT_0_ (Table 2, columns 2 - 3 ≈ column 7) with a residual difference of 0.01 %.

**Table 2:**
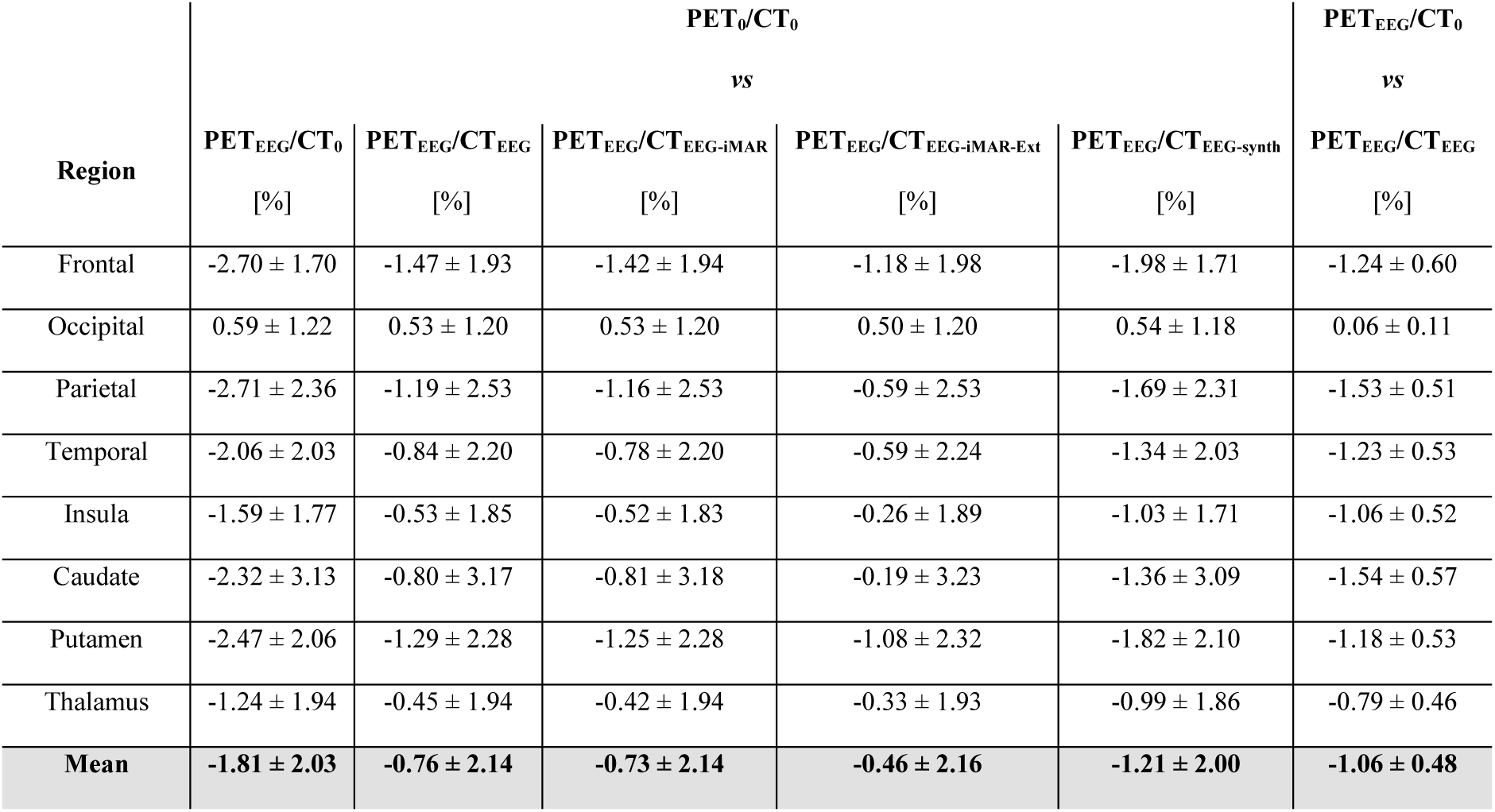
Percentage differences of [^18^F]FDG standard uptake value ratios (SUVR) for different image reconstructions. Columns 2-6 show differences in SUVR compared to the standard reference, i.e., the reconstruction of PET data without EEG using a CT without EEG (PET_0_/CT_0_). This includes reconstructions of PET data with EEG using CT without EEG (PET_EEG_/CT_0_), CT with EEG (PET_EEG_/CT_EEG_), CT with EEG using the iMAR approach without extended HU range (PET_EEG_/CT_EEG-iMAR_), CT with EEG using the iMAR approach and extended HU range (PET_EEG_/CT_EEG-iMAR-Ext_) and CT with EEG electrodes inserted computationally (PET_EEG_/CT_EEG-synth_). See figure 3 for corresponding effects on brain slices. In addition, column 7 shows the comparison between PET data with EEG using CT without EEG (PET_EEG_/CT_0_) and CT with EEG (PET_EEG_/CT_EEG_). This last column approximately matches the difference between columns 2 and 3, i.e., Δ[(PET_EEG_/CT_EEG_) – (PET_EEG_/CT_0_)] when compared to the reference standard. Values correspond to mean ± standard deviation (averaged across subjects and within volumes of interest). iMAR: iterative metal artifact reduction.

| Region | PET<sub>0</sub>/CT<sub>0</sub> |  |  |  |  | PET<sub>EEG</sub>/CT<sub>0</sub> |
| --- | --- | --- | --- | --- | --- | --- |
|  | vs |  |  |  |  | vs |
|  | PET<sub>EEG</sub>/CT<sub>0</sub> | PET<sub>EEG</sub>/CT<sub>EEG</sub> | PET<sub>EEG</sub>/CT<sub>EEG-iMAR</sub> | PET<sub>EEG</sub>/CT<sub>EEG-iMAR-Ext</sub> | PET<sub>EEG</sub>/CT<sub>EEG-synth</sub> | PET<sub>EEG</sub>/CT<sub>EEG</sub> |
|  | [%] | [%] | [%] | [%] | [%] | [%] |
| Frontal | -2.70 ± 1.70 | -1.47 ± 1.93 | -1.42 ± 1.94 | -1.18 ± 1.98 | -1.98 ± 1.71 | -1.24 ± 0.60 |
| Occipital | 0.59 ± 1.22 | 0.53 ± 1.20 | 0.53 ± 1.20 | 0.50 ± 1.20 | 0.54 ± 1.18 | 0.06 ± 0.11 |
| Parietal | -2.71 ± 2.36 | -1.19 ± 2.53 | -1.16 ± 2.53 | -0.59 ± 2.53 | -1.69 ± 2.31 | -1.53 ± 0.51 |
| Temporal | -2.06 ± 2.03 | -0.84 ± 2.20 | -0.78 ± 2.20 | -0.59 ± 2.24 | -1.34 ± 2.03 | -1.23 ± 0.53 |
| Insula | -1.59 ± 1.77 | -0.53 ± 1.85 | -0.52 ± 1.83 | -0.26 ± 1.89 | -1.03 ± 1.71 | -1.06 ± 0.52 |
| Caudate | -2.32 ± 3.13 | -0.80 ± 3.17 | -0.81 ± 3.18 | -0.19 ± 3.23 | -1.36 ± 3.09 | -1.54 ± 0.57 |
| Putamen | -2.47 ± 2.06 | -1.29 ± 2.28 | -1.25 ± 2.28 | -1.08 ± 2.32 | -1.82 ± 2.10 | -1.18 ± 0.53 |
| Thalamus | -1.24 ± 1.94 | -0.45 ± 1.94 | -0.42 ± 1.94 | -0.33 ± 1.93 | -0.99 ± 1.86 | -0.79 ± 0.46 |
| Mean | -1.81 ± 2.03 | -0.76 ± 2.14 | -0.73 ± 2.14 | -0.46 ± 2.16 | -1.21 ± 2.00 | -1.06 ± 0.48 |

**Fig. 3:**
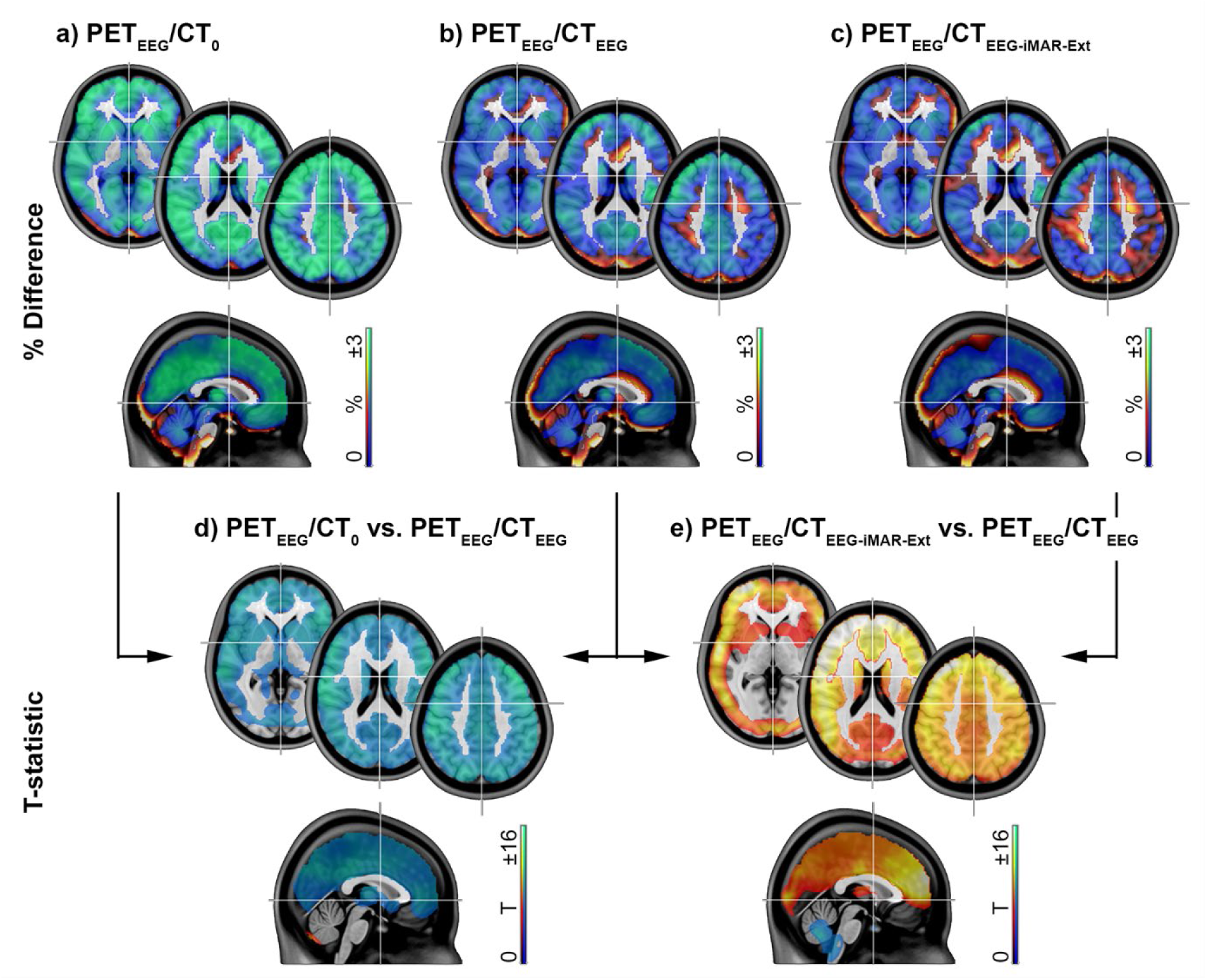
Relative *differences of [^18^F]FDG standard uptake value ratios (SUVR) for different image reconstructions*. **a-c)** Differences in SUVR compared to the standard reference, i.e., the reconstruction of PET data without EEG using a CT without EEG (*PET_0_/CT_0_*). These include reconstructions of PET data with EEG using **a)** CT without EEG (PET_EEG_/CT_0_), **b)** CT with EEG (PET_EEG_/CT_EEG_) and **c)** CT with EEG using the iMAR approach and extended Hounsfield unit range (PET_EEG_/CT_EEG-iMAR-Ext_). **d-e)** Paired, two-sided t-tests comparing PET data (SUVR) with EEG using **d)** CT without vs. with EEG (PET_EEG_/CT_0_ vs. PET_EEG_/CT_EEG_) and **e)** CT with EEG using the iMAR approach and extended Hounsfield units vs. with EEG (PET_EEG_/CT_EEG-iMAR-Ext_ vs. PET_EEG_/CT_EEG_). Statistical tests in **d)** and **e)** are corrected for multiple comparisons at p_FWE_ < 0.05 voxel level. Axial slices are shown at 0, 20 and 45 mm MNI space. Data visualization with MRIcron, arranged in Photoshop.

Overall, the frontal cortex exhibited the lagest differences to the reference (from −2.70 to −1.18 %), whereas approaches performed almost equally across the occipital cortex (from 0.50 to 0.59 %). Although these differences were small in magnitude, the bias was highly consistent across subjects, yielding statistically significant differences between the approaches across almost the entire brain (p_FWE_ <0.05 corrected at voxel level, Figure 3d-e). As expected from the electrode positioning, these differences were least pronounced for the cerebellum and the inferior temporal lobe (Figure 4).

**Fig. 4:**
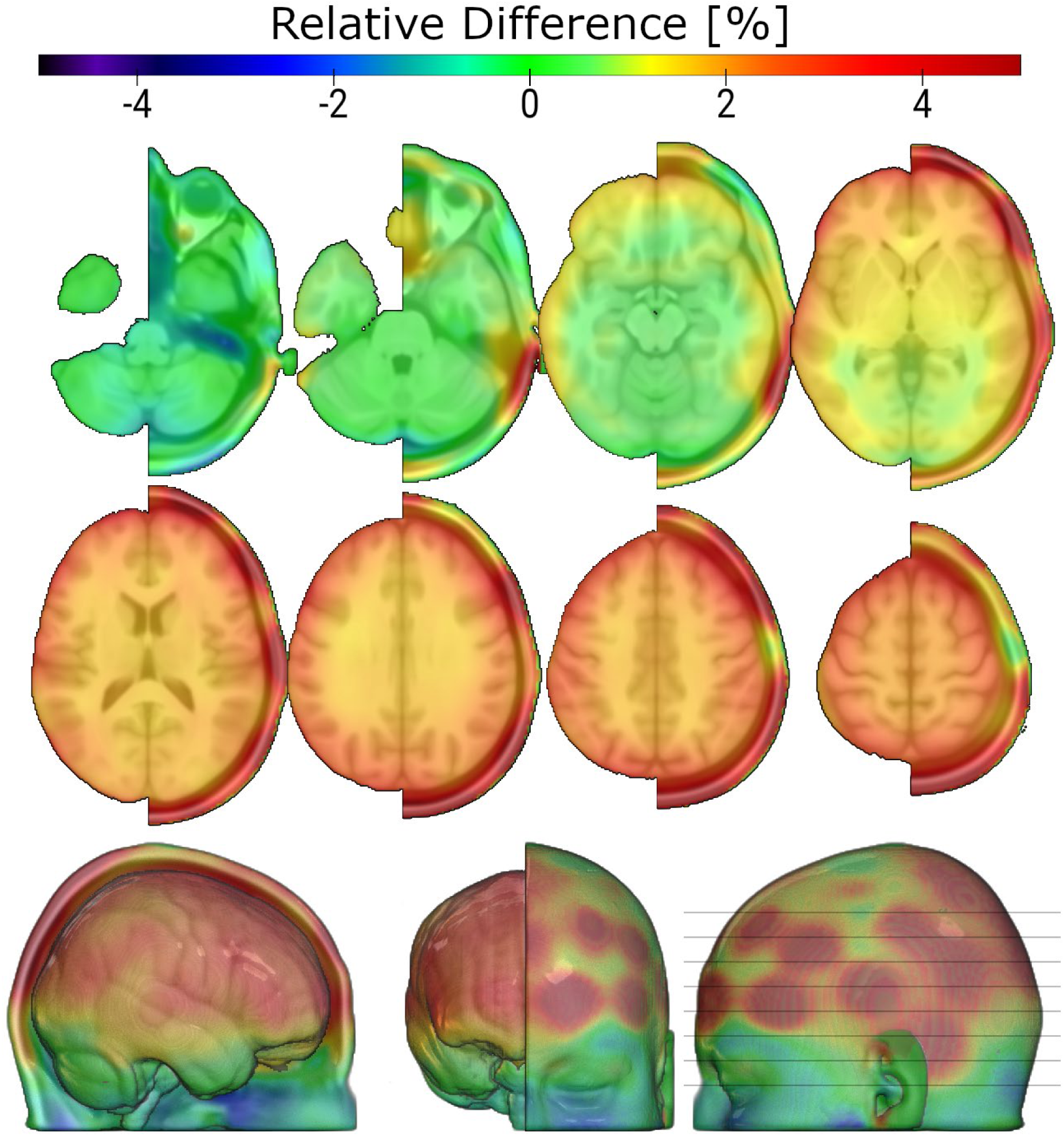
Relative SUVR difference in % across all subjects between PET_EEG_/CT_EEG-iMAR-Ext_ and PET_EEG_/CT_0_. Note the gradient along the z-axis inside of the brain, with the cerebellum and temporal poles being little affected, while regions in planes of many electrodes show larger differences. Axial slices are shown at −45 to 60 mm MNI space in 15 mm intervals. The right hemisphere has been masked to visualize the cortical surface, where electrode positions are less pronounced than on the scalp, but their effects remaining visible. Data visualization with MRIcroGL, arranged in GIMP.

## Discussion

This work evaluates different CT-based strategies for attenuation correction of PET images and metal artifact correction of CTs when EEG electrodes are present during imaging. The CT_EEG_ and CT_EEG-iMAR_ approach, as obtained directly from the scanner, yielded the smallest differences relative to the reference CT_0_ (3.15 ± 3.88 % and −4.02 ± 4.96 %, respectively), while other metal artifact reduction techniques showed stronger bias in AC maps. PET images reconstructed with these different approaches revealed overall small, but highly consistent bias, resulting in statistically significant effects within almost the entire brain in voxel-wise analyses (see Figure 3 d and e). No correction for EEG electrodes (PET_EEG_/CT_0_) performed worst (with relative difference to the reference ranging from −2.71 to 0.59 %), followed by synthetic incorporation of electrodes (PET_EEG_/CT_EEG-synth,_ −1.18 to 0.5 %). Default consideration of EEG in the CT (PET_EEG_/CT_EEG_, −1.47 to 0.53 %) performed similar to the iMAR correction (PET_EEG_/CT_EEG-iMAR_, −1.42 to 0.53 %), and the lowest differences were found with extended HU range (PET_EEG_/CT_EEG-iMAR-Ext_, −1.18 to 0.50 %).

The various MAR techniques were mainly utilized to reduce artifacts in CT images emerging from metallic implants (e.g., dental implants, screws, artificial joints), but no algorithm exists to specifically correct for artifacts of EEG electrodes. This may explain the suboptimal performance of almost all MAR approaches. As iMAR application can be selected in the recording software, it is a convenient solution, that avoids further computations and data handling, but may not be available on all scanner systems.

Incorporation of EEG electrodes in the CT image for attenuation correction (PET_EEG_/CT_EEG_) results in small differences with the reference PET_0_/CT_0_ (−0.76 ± 2.14 %). The performance was similar for the iMAR technique (PET_EEG_/CT_EEG-iMAR_, −0.73 ± 2.14 %), but differences were further reduced when using the extended HU range (PET_EEG_/CT_EEG-iMAR-Ext_, −0.46 ± 2.16 %). Assuming that previous work also employed reconstruction with an extended HU range, this aligns with previous publications showing that MAR techniques provide an optimal solution for PET imaging of the brain [23] and other organs [4]. Detailed inspection of CT images shows that iMAR effectively removes streak artifacts, which emerge from x-rays attenuated and scattered by EEG electrodes (see Figure 2). However, a CT scan with EEG electrodes present may not always be available, particularly when using a PET/MR scanner. We therefore tested whether the artificial incorporation of electrodes into the CT_0_ image would prove sufficient to account for attenuation of the EEG (PET_EEG_/CT_EEG-synth_). This resulted in a reduced bias (1.21 ± 2.00 %) as compared to PET_EEG_/CT_0_ (−1.81 ± 2.03 %), but did not perform alike to PET_EEG_/CT_EEG_. We speculate that capturing additional differences such as cables and EEG gel, as well as more sophisticated inclusion of electrodes may yield a better outcome.

The complete negligence of electrodes in attenuation of PET images (PET_EEG_/CT_0_) resulted in the most pronounced, but still small (<3%) bias compared to the reference. This might explain the lack of observed differences when evaluated visually [24], which is also unlikely to affect clinical diagnosis [1, 23]. Nevertheless, statistical differences were observed in the current and already previous work [23] but not reported for other studies [25], although individual, local differences can reach 10-15 % [25, 26]. Albeit this difference might not affect within-scan assessment of changes [26], it needs to be considered for absolute quantification and when combining data from different acquisition protocols, i.e., with and without EEG, to avoid introduction of systematic bias. Furthermore, the effects observed in the current work exhibited regional variability, with the frontal cortex showing the largest differences across all correction approaches, which was up to four-fold higher than for other regions. This may affect the estimation of molecular covariance as such approaches investigate interregional correlations across time or subjects [27].

The comparisons in this work were made to a reference without EEG (PET_0_/CT_0_). Although it was acquired about 10 min later than images with EEG, detailed comparisons showed negligible differences (residual difference < 0.01%), indicating that this temporal acquisition difference in which FDG decays and physiological changes in tracer uptake could occur, did not drive the results. Another limitation is the use of only 32 EEG channels, since more sophisticated systems employ up to 256 leads. However, already this limited number of electrodes showed a consistent bias throughout the brain. Future work should therefore investigate the extent to which this bias scales with larger numbers of electrodes.

In conclusion, EEG electrodes induce a small, but consistent bias in PET images. This should be considered, specifically when pooling data with and without simultaneous EEG and for interregional assessments such as molecular connectivity. The effect can, however, be reduced by incorporating electrodes into the attenuation correction.

## Acknowledgements

We thank the graduated team members and the diploma students of the Neuroimaging Labs (NIL, head R. Lanzenberger) as well as the clinical colleagues from the Department of Psychiatry and Psychotherapy and radiotechnologists from the Department of Nuclear Medicine for their valuable support. Special thanks go to David Gomola for administrative work, Godber M Godbersen, Clemens Schmidt and Elisa Briem for clinical expertise, and David Nolz for computational support.

The scientific project was performed with the support of the Research Platform Medical Imaging (RPMI) of the Medical University of Vienna.

## Statements and Declarations

## Funding

This research was funded in whole, or in part, by the Austrian Science Fund (FWF) [grant DOIs: 10.55776/KLI1006 and 10.55776/PAT6608924; PI: R. Lanzenberger]. G Dörl and C Milz are recipients of a DOC fellowship of the Austrian Academy of Sciences at the Department of Psychiatry and Psychotherapy, Medical University of Vienna.

For open access purposes, the author has applied a CC BY public copyright license to any author accepted manuscript version arising from this submission.

## Author Contributions

**Data Acquisition** PS, MP, GD, CM, SG, BE, MR, MM, PF, KA, AH

**Data Curation and Analysis** PS, MP, CM, AH

**Writing – Initial Draft** PS, MP, AH

**Writing – Review, Comments and Editing** PS, MP, GD, CM, SG, BE, MR, MM, PF, KA, LN, SR, MH, RL, AH

**Methodology and Resources** LN, SR, MH, RL, AH

**Funding** RL, GD, CM

## Data Availability

Raw data will not be publicly available due to reasons of data protection. Processed data and custom code can be obtained from the corresponding author with a data-sharing agreement, approved by the departments of legal affairs and data clearing of the Medical University of Vienna.

## Ethics Approval

The study was approved by the Ethics Committee of the Medical University of Vienna (ethics number: 1642/2022) and all procedures were carried out in accordance with the Declaration of Helsinki.

## Consent to Participate and Publish

After detailed explanation of the study protocol, all subjects gave written informed consent. All subjects were insured and reimbursed for their participation. Consent to publish is not applicable.

## Abbreviations and Acronyms

AC: attenuation correction
CT: computed tomography (scan)
CT_0_: without EEG electrodes present
CT_EEG_: with EEG electrodes present
CT_EEG-ANRF_: with application of ANRF
CT_EEG-iMAR_: with application of iMAR (see MAR)
CT_EEG-iMAR-Ext_: with extended Hounsfield unit range
CT_EEG-MDT_: with application of MDT (see MAR)
CT_EEG-NMAR_: with application of NMAR (see MAR)
CT_EEG-RMAR_: with application of RMAR (see MAR)
CT_EEG-synth_: CT_0_ with digitally (“synthetically”) added EEG electrodes
EEG: electroencephalogram
([^18^F])FDG: ([^18^F])fluorodeoxyglucose
HU: Hounsfield units
ΔHU_min_: minimal increments of HU in an image, i.e., HU resolution
LAC: linear attenuation coefficient
MAR: metal artefact reduction
ANRF: 2-D adaptive noise-removal filtering
iMAR: iterative MAR (proprietary)
MDT: metal deletion technique
NMAR: normalized MAR
RMAR: refined MAR
PET: positron emission tomography (scan)
PET_0_: without EEG electrodes present
PET_EEG_: with EEG electrodes present
PET_x_/CT_y_: PET_x_ reconstructed using CT_y_ for attenuation correction
STD: standard deviation
SUVR: standard uptake value ratio
T: time
VOI: volume of interest

